# Evoking highly focal percepts in the fingertips through targeted stimulation of sulcal regions of the brain for sensory restoration

**DOI:** 10.1101/2020.11.06.20217372

**Authors:** Santosh Chandrasekaran, Stephan Bickel, Jose L Herrero, Joo-won Kim, Noah Markowitz, Elizabeth Espinal, Nikunj A Bhagat, Richard Ramdeo, Junqian Xu, Matthew F Glasser, Chad E Bouton, Ashesh D Mehta

## Abstract

Paralysis and neuropathy, affecting millions of people worldwide, can be accompanied by a significant loss of somatosensation. With tactile sensation being central to achieving dexterous movement, brain-computer interface (BCI) researchers have explored the use of intracortical electrical stimulation to restore sensation to the hand. However, current approaches have been restricted to stimulating the gyral areas of the brain while functional imaging suggests that the representation of fingertips lie predominantly in the sulcal regions. Here we show, for the first time, highly focal percepts can be evoked in the fingertips of the hand through electrical stimulation of the sulcal areas of the brain. To this end, we mapped and compared sensations elicited in the hand by stimulating both gyral and sulcal areas of the human primary somatosensory cortex (S1). Two participants with intractable epilepsy were implanted with stereoelectroencephalography (SEEG) and high-density electrocorticography (HD-ECoG) electrodes in S1 guided by high-resolution functional imaging. Using myelin content and cortical thickness maps developed by the Human Connectome Project, we elucidated the specific sub-regions of S1 where focal percepts were evoked. Within-participant comparisons showed that sulcal stimulation using SEEG electrodes evoked percepts that are significantly more focal, with 80% less area of spread (p=0.02) and localized to the fingertips more often than in gyral stimulation via HD-ECoG electrodes. Finally, sulcal locations exhibiting repeated modulation patterns of high-frequency neural activity during mechanical tactile stimulation of the hand showed the same somatotopic correspondence as sulcal stimulation. These findings show that minimally-invasive sulcal stimulation could lead to a clinically viable approach to restoring sensation in those living with sensory impairment.

**Significance:** Intracortical or cortical surface stimulation of the primary somatosensory cortex (S1) offers the promise of restoring somatotopically-relevant sensation in people with sensory impairment. However, evoking percepts in the fingertips has been challenging as their representation has been shown to be predominantly located within sulcal regions of S1 – inaccessible by these stimulation approaches. We evoked highly focal percepts in the fingertips of the hand by stimulating the sulcal regions of S1 in people with intractable epilepsy using stereoelectroencephalography (SEEG) depth electrodes. Sensory percepts in the fingertips were more focal and more frequently evoked by SEEG electrodes than by high-density electrocorticography (HD-ECoG) grids evidenced by within-participant comparisons. Our results suggest that fingertip representations are more readily targeted within the sulcal regions. SEEG electrodes potentially offer a clinically viable approach to access the sulcal regions for sensory neuroprostheses that can aid dexterous motor control.

## Introduction

Over 5 million people are living with paralysis in the United States alone (1). The two leading causes of paralysis, namely stroke and spinal cord injury (SCI), also result in sensory impairment of the upper limb in up to 12% (2) and 54% (3) of individuals, respectively. Meanwhile, of about 422 million people worldwide with diabetes mellitus (4), up to 64% can experience peripheral neuropathy in their hands leading to significant impairment of the sense of touch (5). Such loss of sensation critically impairs the ability to perform dexterous manipulation of objects (6, 7). Intracortical brain-computer interfaces (BCI) have shown tremendous success in decoding intended movements from neural activity recorded in the primary motor cortex (8, 9) and subsequently, restoring motor control of their own hand in people with tetraplegia (10). However, this significant progress in neurorehabilitation is often hampered by the lack of tactile feedback. Without somatosensation, users of BCI systems almost entirely rely on visual feedback while interacting with objects precluding fine motor control, such as manipulation of small objects, inability to detect object contact to transition from reaching to grasping, modifying grasp strength to prevent slipping, or interacting with objects outside the line of sight.

Recently, tactile percepts in the hand have been evoked in humans through intracortical microstimulation using microelectrode arrays (11–13) or cortical surface stimulation using electrocorticography (ECoG) grids (14–16) in the primary somatosensory cortex (S1). Although such artificial sensory feedback help improve the user performance with a BCI system (17), focal percepts in fingertips that would be critical for dexterous manipulations have been difficult to achieve. In a recent study, targeting fingertip representations required extensive intraoperative mapping using mechanical stimulation at the periphery (13) relying on spared neural pathways which may not be feasible in many patients with SCI. A primary reason for the inability to reliably evoke fingertip percepts could be that cortical stimulation in these studies has been restricted to the gyral areas of S1, namely the postcentral gyrus.

Functional magnetic resonance imaging (fMRI) of human S1 has shown individual digit representations to occur in the central and postcentral sulcus in addition to the postcentral gyrus covering the cytoarchitectonically distinct cortical areas 3b, 1 and 2 (18–21). As observed in non-human primates (22–24), studies suggest a mirror-reversal of phalange representation to exist at the area 3b/1 border located in the central sulcus close to the crown of the postcentral gyrus. This places the proximal phalanges close to that border while more distal phalanges, including the fingertips, occur towards the area 3a/3b border located at the base of the central sulcus (18, 25) and in the posterior regions of area 1 located over the postcentral gyrus (18, 25, 26). This would be consistent with the observations from a recent human somatosensory mapping study (27). However, other studies show representation of distal phalanges along the posterior wall of the central sulcus but closer to the 3b/1 border (28, 29). Thus, it is still unclear how the representation of fingertips is distributed across the central sulcus and postcentral gyrus (30).

Recent advances in stereotactic placement of depth electrodes, as in stereoelectroencephalography (SEEG), increasingly provide reliable access to deeper cortical and subcortical targets in the brain (31). These electrodes are increasingly used in the clinic for seizure onset localization in patients with medically refractory epilepsy (32). In addition, SEEG electrodes have been used to map and document the sensory percepts evoked while stimulating the human parietal (33) and insular cortices (34). Comparing separate cases involving SEEG and ECoG implantations, SEEG electrodes have been shown to be a clinically useful alternative for electrical brain stimulation (EBS) to map eloquent cortical areas (35, 36). In fact, a recent study showed that SEEG-mediated EBS could identify sensorimotor areas with high accuracy and specificity (37). Moreover, implantation procedures for SEEG electrodes are minimally invasive (∼2mm craniostomy) with lower rates of infection (38) compared to subdural strip and ECoG electrodes which require a craniotomy several centimeters wide (39).

In this first-in-human study, we explored the representation of the hand in the sulcal regions of S1 using SEEG electrodes. We implanted both SEEG and HD-ECoG electrodes in two patients with intractable epilepsy. A within-participant comparison of the percepts evoked by the two electrode types allowed us to map the hand representations in both the gyral and sulcal areas of S1 and compare the corresponding evoked percepts. Electrode implantation was guided by high-resolution functional magnetic resonance imaging (fMRI) obtained during a finger-tapping task analyzed using the Human Connectome Project (HCP) processing pipelines. T1-weighted (T1w) and T2-weighted (T2w) structural images provided the T1w/T2w-based myelin content and cortical thickness maps which enabled atlas-based parcellation of the cortical areas, somatotopic subregions, and sub-areas of the sensorimotor cortex (40) further informing electrode implantation. Upon administering intracortical direct electrical stimulation using either SEEG or HD-ECoG electrodes, the participants reported sensory percepts that were localized to the contralateral arm, hand, and even fingertips. Strikingly, we observed that tactile percepts evoked by SEEG electrodes were much more focused in their spatial extent. Furthermore, the percepts evoked by SEEG electrodes tended to be in and around the fingertips more often. SEEG-mediated recording of neural activity evoked upon mechanical tactile stimulation enabled precise localization of cortical regions involved in processing of sensory information from specific finger and palm regions.

These results demonstrate that stimulation through SEEG electrodes can effectively access the sulcal regions of S1 and activate fingertip representations. Combined with their minimally invasive implantation procedure, SEEG electrodes can be an effective and reliable approach to evoke focal percepts in the hand and fingers that are functionally relevant for people with tetraplegia. Furthermore, they can be an effective tool for passive mapping of the brain for clinical purposes.

## Methods

### Participants

Two patients undergoing pre-operative seizure monitoring for surgical treatment of intractable epilepsy took part in this study. The patients were implanted with either HD-ECoG grids and/or SEEG electrode leads. The decisions regarding whether to implant, the electrode targets, and the duration for implantation were based entirely on clinical grounds without reference to this investigation. Patients were informed that participation in this study would not alter their clinical treatment, and that they could withdraw from the study at any time without jeopardizing their clinical care. All procedures and experiments were approved by the Northwell Institutional Review Board and participants provided informed consent prior to enrollment into the study.

### Imaging

Participants were scanned on a 3T MRI scanner (Skyra, Siemens, Germany) with a 32-channel head coil. HCP-like structural and functional MRI were acquired: T1-weighted (T1w) 3D MPRAGE sequence, 0.8 mm isotropic resolution, TR/TE/TI = 2400/2.07/1000 ms, flip angle = 8 degree, in-plane under-sampling (GRAPPA) = 2, acquisition time 7 min; T2-weighted (T2w) 3D turbo spin echo (SPACE) sequence, 0.8 mm isotropic resolution, in-plane under-sampling (GRAPPA) = 2, TR/TE = 3200/564 ms, acquisition time 6.75 min; task fMRI using the CMRR implementation of multiband gradient echo echo-planar imaging (EPI) sequence (41), 2.1 mm isotropic resolution, 70 slices with a multiband factor of 7 (42), FOV 228 mm × 228 mm, matrix size 108 × 108, phase partial Fourier 7/8, TR/TE = 1000/35 ms, flip angle = 60 degree, phase encoding direction = anterior-posterior (A-P), echo spacing = 0.68 ms, 240 volumes in 4 min; and a pair of reversed polarity (A-P / P-A) spin echo EPI field mapping acquisitions with matched echo train length and echo spacing to the fMRI acquisition. The task was button-pressing on the PST button response unit (Psychology Software Tools, Sharpsburg, PA, USA) using a single finger (wrist restrained with strap on the button response unit and neighboring fingers taped down with medical tape), repeating 6 times of 20-second off (resting with cue of a blank dark screen) and 20-second on (tapping with continuous video cue of the same finger motion presented from a projector screen). Participant 1 performed three repetitions of the task for each of thumb, index, and little fingers (phase-encoding direction A->P) while participant 2 performed two repetitions for each of thumb, index, and middle fingers (phase-encoding directions A->P and P->A). The MRI preprocessing began with the HCP minimal preprocessing pipelines version 3.27 (43) including, motion correction, distortion correction, cortical surface reconstruction and subcortical segmentation, generation of T1w/T2w-based myelin content and cortical thickness maps, transformation of the fMRI data to MNI and CIFTI grayordinate standard spaces using folding-based registration with MSMSulc (44, 45), and 2 mm FWHM surface and subcortical parcel constrained smoothing for regularization. The fMRI data were cleaned of spatially specific structured noise using the HCP’s multi-run (version 4.0) ICA-FIX (46–48) for multi fMRI (multiple finger tasks) and linear trends without regressing out motion parameters. Somatotopic functional responses were estimated (first-level for participant 1 and second-level fixed-effect averaging of the two phase-encoding directions for participant 2) using a generalized linear model (GLM)-based fMRI analysis (49) on the grayordinate data space for each finger.

### Electrode localization

The SEEG electrodes (PMT Corporation, Chanhassen, MN, USA) consisted of 16 contacts, cylinders with 2 mm length, 0.8 mm diameter, and 4.43 mm spacing (center to center). The HD-ECoG grids (PMT Corporation) consisted of 2 mm diameter flat contacts, in participant 1 with an 8×8 arrangement with 5 mm spacing (center to center) and in participant 2 with 16×16 contact arrangement with 4 mm spacing. Since both patients had clinical indications that required mapping of the sensorimotor cortex, task-based fMRI activation maps were used to guide electrode placement.

For digital localization of the electrodes, we used the freely available iElvis toolbox, available at https://github.com/iELVis/ (50). Briefly, the electrodes were manually localized using the software BioImage Suite (http://www.bioimagesuite.org) on a postimplant CT which was co-registered using an affine transformation (6 degrees-of-freedom FLIRT; www.fmrib.ox.ac.uk/fsl) to a pre-implantation 3T high-resolution T1w MRI. We used the FreeSurfer (51) output from the HCP minimal processing pipeline (40) to obtain the pial surface. The subdural HD-ECoG electrodes were projected to the smoothed pial surface. The smoothed pial surface, also called the outer smoothed surface, is generated by Freesurfer and wraps tightly around the gyral surfaces of the pial layer while bridging over the sulci. No correction was applied to SEEG electrode coordinates.

To visualize the fMRI activation maps and the electrodes simultaneously, we used HCP Connectome Workbench. Before importing the electrode coordinates into Workbench, we applied a RAS coordinate offset as follows –

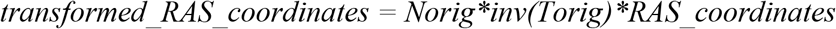

where the transformation matrices *Norig* is obtained by *mri_info --vox2ras [subject]/mri/orig*.*mgz* and *Torig* is obtained by *mri_info --vox2ras-tkr [subject]/mri/orig*.*mgz*

The transformed coordinates were then imported as foci using the T1w surfaces into a developmental version of Connectome Workbench.

### Electrical brain stimulation (EBS)

Intracranial EBS is routinely used clinically to identify eloquent cortex to be spared from surgical resection. For this study, we used routine EBS parameters with a S12D Grass cortical stimulator (Grass Technologies, Pleasanton, CA). We used bipolar pairs of electrodes for stimulation and delivered current-regulated, symmetric alternating square-wave pulses with 0.2 ms width per phase, at 20 or 50 Hz, with stimulation amplitudes between 0.5–6 mA, for 0.5–2 seconds while the participant was quietly resting and asked to report the occurrence of any sensation. Stimulation amplitude was slowly increased until a percept was elicited, or after-discharges occurred up to a maximal amplitude of 6 mA. If a sensation on the hand was reported, the participant was asked to draw the affected area on a schematic of a hand. The sensations were described as “tingling” or “sensation of electricity”. Without informing the participant, sham trials (0 mA stimulation) were intermixed with real stimulation trials. Intracranial EEG was acquired continuously using a clinical recording system (XLTEK, Natus Medical) at 512 Hz or 1 kHz and monitored across all implanted electrodes for the presence of after-discharges and seizures. No seizures were caused during stimulation of the areas reported here.

### Analysis of sensory percepts

For this study, we focused our analysis to the sensory percepts localized to the hand and wrist. To digitize the participant responses, we used a script custom-written in MATLAB to redraw the participant drawings on a computer. Surface areas of the digitized percepts were then calculated and used for all further analysis.

### Recording of neural activity

In addition to the clinical recording system, neural activity was recorded for participant 2 using a Neuroport System (Blackrock Microsystems, Salt Lake, Utah) with a sampling rate of 10 kHz while performing mechanical stimulation of the fingertips of their left hand. We used a vonFrey filament (TouchTest Sensory Probes) of evaluator size, 3.61 (0.4 g). An electrode located in soft tissue lacking neural activity was used as the system ground. Subsequent analysis involved multiple steps to extract information regarding power modulation in different frequency bands. Signals from neighboring channels were subtracted in software to provide bipolar data with reduced noise. Non-overlapping Blackman windows of 200 ms in length were applied to the data, followed by a short Fast Fourier Transform (sFFT) for each window (with a resulting frequency resolution of 5 Hz). The amplitude information at each frequency was then integrated across pre-selected frequency bands as follows: 0–10, 10–15, 15– 30, 30–100, 100–500, and 500–5000 Hz. These integrated amplitude features for all bipolar recordings were standardized by subtracting their mean and dividing by their standard deviation across the entire task. For the analysis in this study, we chose neural activity in the high gamma or 100-500 Hz frequency band. Epochs aligned with each visual cue (animated hand) presented to the participant during each task were created, starting at the cue onset, and extending to 400 ms after the cue offset. All aligned trials for each epoch (cue) type were averaged to form a composite temporal response. To quantify the degree of repeatability, the mean correlation coefficient (MCC) was computed by averaging the correlation coefficients obtained for temporal responses for each trial with respect to the composite.

## Results

Both study participants were first implanted with SEEG leads, subsequently replaced by a HD-ECoG grid, for extraoperative monitoring of neural activity to localize the epileptogenic zone. During routine clinical intracranial EBS using either of these electrodes, participants were asked to report the sensory percepts that were evoked. For this study, we focused on the sensory percepts localized to the hand and wrist. Stimulation amplitudes that evoked sensations in the hand area ranged from 0.5–6 mA with stimulation frequencies of either 20 or 50 Hz, and a pulse width of 200 µs. After each stimulation trial (lasting 0.5–2 s), the evoked percept was reported by the participant and was recorded by the experimenter. A total of 43 electrode pairs (12 SEEG and 31 HD-ECoG electrodes) across the two participants evoked at least one sensory percept in the contralateral hand or arm. Other electrodes evoked percepts that were localized to more proximal areas of the arm or even perceived bilaterally. These were not included in the analysis performed during this study.

### HD-ECoG and SEEG electrodes provide access to different cortical areas

To locate the representation of the fingers in the primary somatosensory cortex (Figure 1 insets), the participants performed button-press tasks using thumb (D1, red), index (D2, green), and little finger (D5, blue, participant 1) or middle finger (D3, blue, participant 2) during functional imaging. The fMRI activation results were thresholded to optimize the visualization of topological features in area 3b (Figure 1, right panels), located on the posterior wall of the central sulcus, and overlaid (from top to bottom without transparency: red, green, and blue) on group average cortical areal maps (40). Overlapping activation in area 4 (anterior wall of the central sulcus) and 3a (nadir of the central sulcus) are evident (most of the blue and a large portion of the green are hidden beneath the red color) in both participants. A topologically meaningful representation of D1, D2, and D3/D5 can be observed in the lateral-medial axis in area 3b in both participants. Interestingly, representation even more medial to the D3/D5 representation can be observed in D1 (red) and D2 (green) tasks. Less consistent individual digit representations were observed in area 1 (postcentral gyrus), except for an overlapping activation in the lateral location of D1 for all finger tasks. Even less consistent or appreciable activations exist in area 2 at the chosen threshold level.

**Figure 1.**
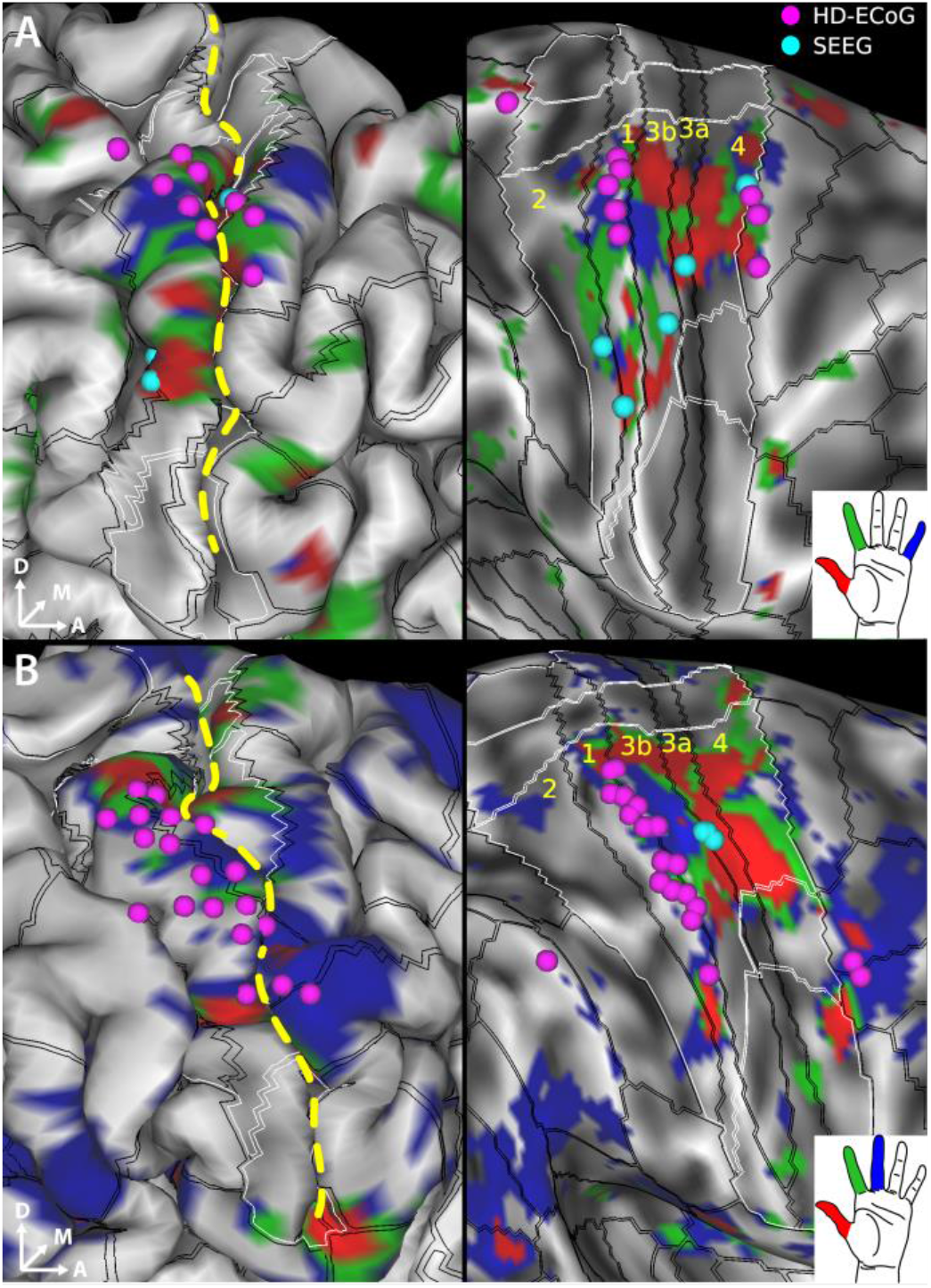
fMRI activation maps for individual digits and electrode sites evoking sensory percepts in the hand region upon electrical stimulation. Sensorimotor cortex activation map shown in **A**. for digits D1 (red), D2 (green) and D5 (blue) in participant 1 and in **B**. for digits D1 (red), D2 (green) and D3 (blue) in participant 2. SEEG (cyan spheres) and HD-ECoG (fuchsia spheres) electrodes that evoked at least one sensory percept in the hand area are overlaid over the pial surface (left panels) and over the “very-inflated” representation of the cortical surface (right panels). The black lines overlaid on the cortical surface delineate the cortical areal boundaries outside of sensorimotor cortex, and sensorimotor subregional boundaries inside sensorimotor cortex as derived from the HCP S1200 group average parcellation (40) based on myelin and cortical thickness maps and are labeled in yellow letters. The white lines demarcate the somatotopic sensorimotor subregions based on resting state and task-based fMRI. The sensorimotor subareas are denoted by the intersection of the black and white boundaries. Gray shading denotes curvature of the cortical surface. Dashed yellow line denotes the central sulcus.

We were interested in further elucidating the digit representations in the sulcal and gyral areas of S1. Specifically, we wanted to determine whether electrical stimulation using SEEG depth electrodes can effectively target these digit representations in the sulcal areas of S1. Overlaying the electrodes that evoked sensations in the hand on top of the fMRI maps shows that the HD-ECoG electrodes (fuschia spheres in Fig. 1) appear to cluster in area 1 which covers the apical surface of the postcentral gyrus. Meanwhile, the SEEG electrodes (cyan spheres in Figure 1) that evoked sensory percepts in the hand, when projected to the cortical surface, localize predominantly to the areas 3a and 3b of S1 which cover the nadir and posterior wall of the central sulcus, respectively. Figure S1 demonstrates that the group average cortical areal definitions align well with the individual subject cortical myelin and thickness maps.

### Fingertip percepts evoked by SEEG electrodes

We characterized the sensory percepts evoked following direct electrical stimulation of S1 using SEEG or HD-ECoG electrodes. In both participants, electrical stimulation was gradually ramped up until percept threshold was reached, at which point the participants described what they perceived. We observed that the percepts evoked by SEEG stimulation tended to be highly focal, often restricted to within a single segment of a finger, and often at the fingertips. The SEEG electrodes that evoke these percepts were predominantly located near the anterior wall of the postcentral gyrus (Figure 2, panels A and C). Meanwhile, the percepts evoked by HD-ECoG electrodes often extended over multiple segments of a digit or multiple digits (Figure 2, panels B and D). We observed a paucity in percepts restricted to fingertips alone when stimulating using HD-ECoG electrodes. As expected, the evoked percepts exhibit a somatotopical organization of the hand, with thumb percepts evoked by electrodes that were more laterally located, while percepts in the index and middle fingers and the wrist were evoked by electrodes that were more dorsal (Figure 2D).

**Figure 2.**
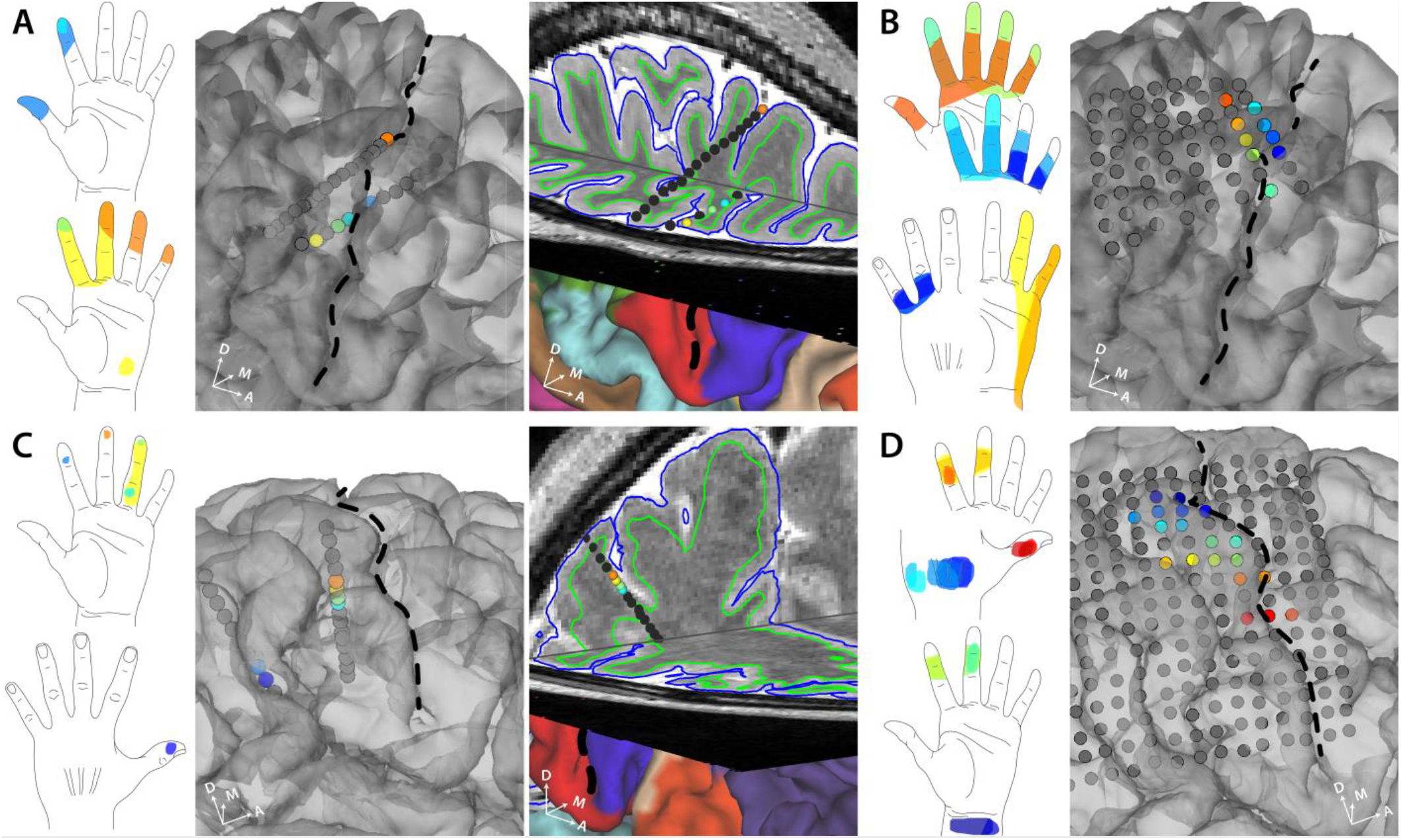
Self-reported sensory percepts in the hand upon stimulation via SEEG or HD-ECoG electrodes. **A**. Sensory percepts reported by participant 1 upon stimulation through SEEG electrodes. The color of each electrode matches the color of the corresponding percept evoked. The third panel shows a 3D brain slice showing the same SEEG electrodes. **B**. Sensory percepts reported by participant 1 upon stimulation through HD-ECoG electrodes. **C**. Sensory percepts reported by participant 2 upon stimulation through SEEG electrodes. The color of each electrode matches the color of the corresponding percept evoked. The third panel shows a 3D brain slice showing the same SEEG electrodes **D**. Sensory percepts reported by participant 2 upon stimulation through HD-ECoG electrodes. Black dashed line denotes the central sulcus.

### Focal and distal percepts evoked by SEEG electrodes

We characterized and compared the percepts evoked by SEEG depth and HD-ECoG grid electrodes. Comparing the areas enclosed by the sensory percepts showed that SEEG electrodes evoked percepts that were significantly smaller than those evoked by HD-ECoG electrodes (Figure 3A, p=0.02, Wilcoxon ranksum test, χ^2^ = 5.57). This suggests that the sensory percepts evoked by SEEG electrodes tend to be more focal in their spatial spread.

**Figure 3.**
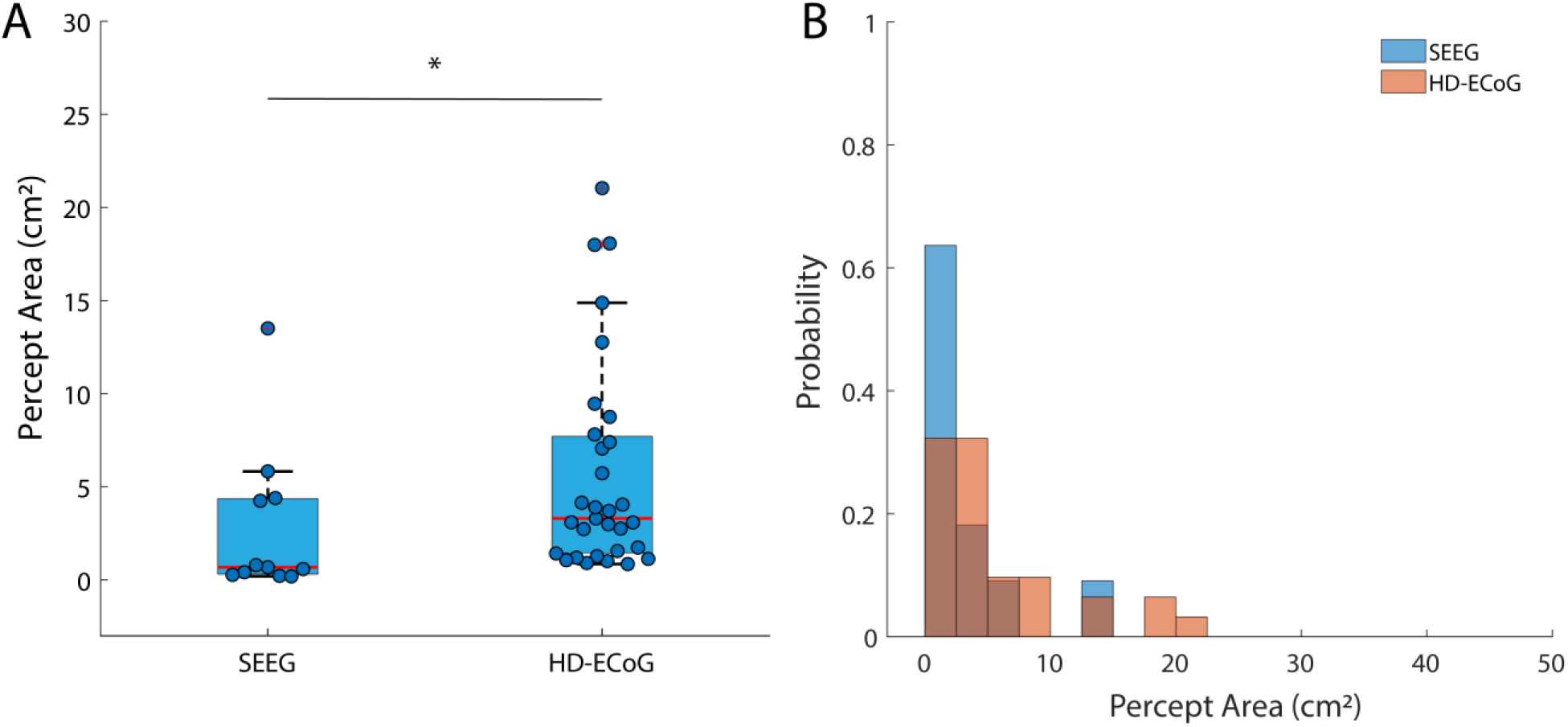
Sensory percepts evoked by SEEG electrodes tend to be more focal in fingers. **A**. Boxplot showing the distribution of the areas covered by sensory percepts evoked by SEEG and HD-ECoG electrodes. **\*** denotes significance in a Wilcoxon ranksum test, χ2 = 5.57; p=0.02. **B**. Histograms showing frequency of occurrence of percepts of different sizes for SEEG (blue) and HD-ECoG electrodes (orange). The two distributions are significantly different in a Komolgorov-Smirnov test, p<<0.01.

Comparing the probability of occurrence of percepts of different sizes also showed significant skew towards percepts with smaller area in case of SEEG electrodes (Figure 3B, p<<0.01, Komolgorov-Smirnov test).

To evaluate if there was a difference in the location of the evoked percepts between the two types of electrodes, we determined the probability that an evoked percept covered a particular area of the hand. The resulting heatmap shows that percepts that spread over the fingertips constituted up to 25-30% of those evoked by SEEG electrodes (Figure 4A) compared to only 10% of those evoked by the HD-ECoG electrodes. The most represented region of the hand in case of percepts evoked by HD-ECoG electrodes were the middle phalanges of the fingers and the ulnar side of the hand (up to 30%, Figure 4B).

**Figure 4.**
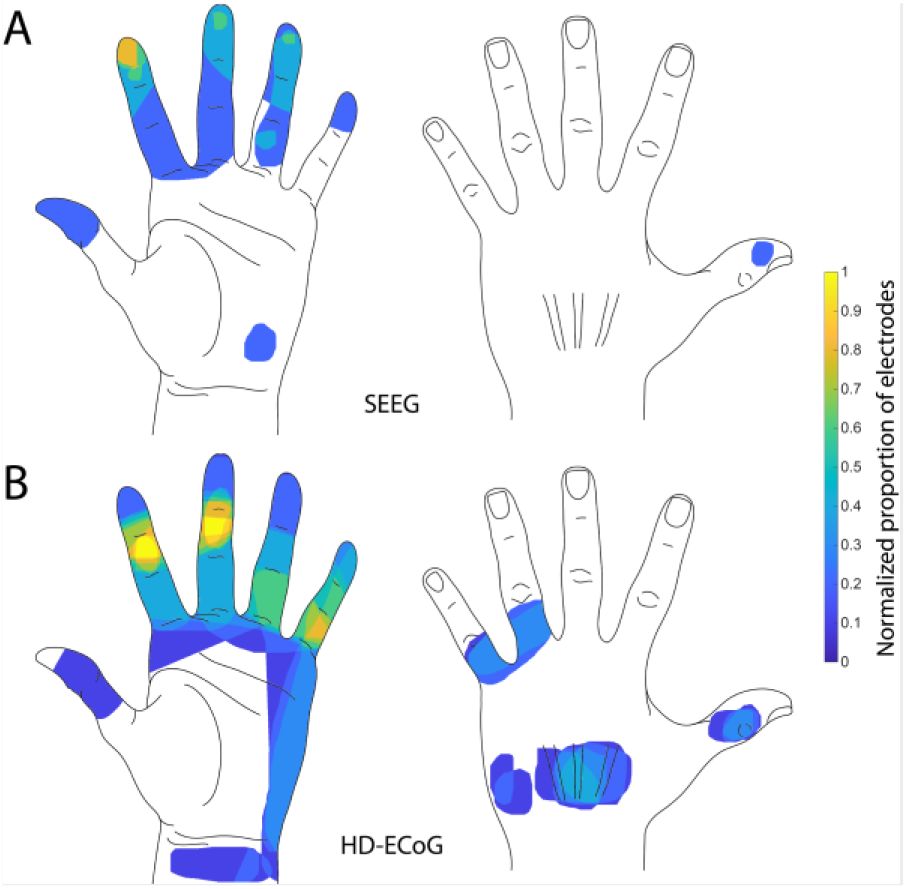
Heatmap of evoked percepts. **A**. Heatmap shows frequency of any region of the hand being part of a sensory percept evoked by SEEG electrodes pooled from both patients. Heatmap shows frequency of any region of the hand being part of a sensory percept evoked by HD-ECoG electrodes pooled from both patients. Electrode numbers were normalized to the maximum number of electrodes that evoked a percept in the same region (n = 5 for SEEG; n = 10 for HD-ECoG).

### Cortical activity recorded during mechanical tactile stimuli

While implanted with SEEG electrodes, Participant 2 also received mechanical tactile stimulation of the thumb, index and middle finger pads and the resultant neural activity in S1 was recorded. We aimed to further confirm the previously identified sulcal locations were involved in fingertip tactile sensation. We identified electrodes from which neural activity were recorded that showed a high degree of repeatability (mean correlation coefficient, r ≥ 0.6) among features in the high gamma band (100–500 Hz) across repeated cycles of mechanical stimulation, i.e., tapping of the fingertips. We show that SEEG electrodes recording stimulus evoked somatosensory activity were spatially clustered (Figure 5A). Interestingly, the SEEG electrodes that evoked a sensory percept in the hand overlapped or were located close to the SEEG electrodes that were activated by tactile stimulation of the fingertips (Figure 5B).

**Figure 5.**
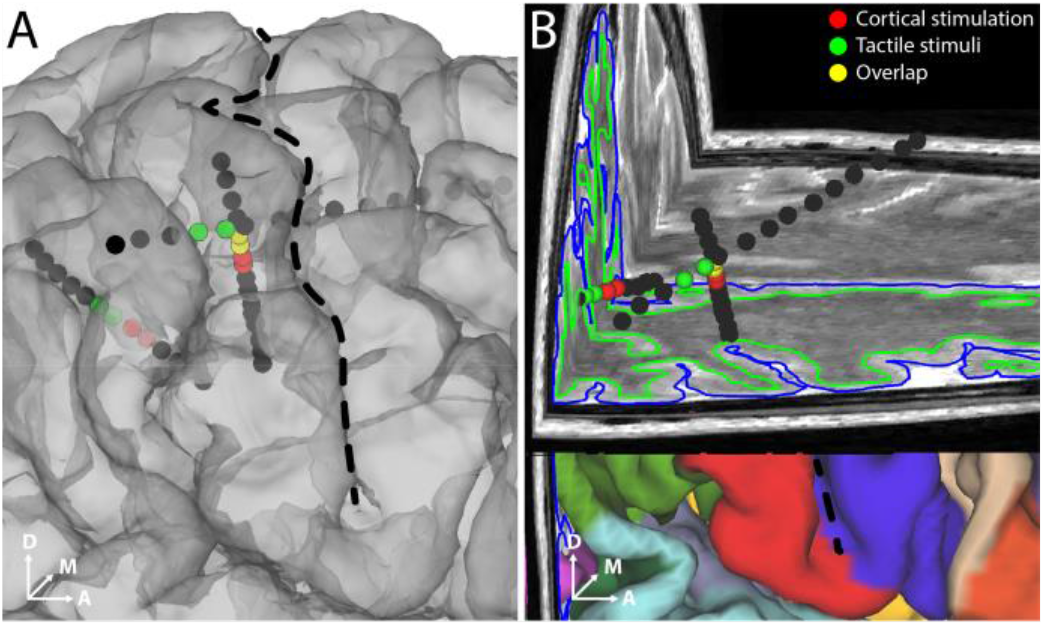
Recorded neural activity correlated with tactile stimulus of fingertips in participant 2. Electrodes that showed high degree of repeatability (r>0.6) of features across mechanical tactile stimulation cycles are shown (green spheres). The electrodes that evoked a percept in the hand area are also shown (red spheres). Overlapping electrodes are shown in yellow. Black dashed line denotes the central sulcus.

## Discussion

In this study, we demonstrate that sulcal stimulation in human S1 evokes sensory percepts localized to the fingertips more often than gyral stimulation. SEEG electrodes provided an effective way to deliver intracortical electrical stimulation to sulcal regions of S1. Using these electrodes, in two participants, we were able to evoke sensory percepts restricted to single segments of a digit, including fingertips and more focal than those evoked by HD-ECoG electrodes. We were also able to record neural correlates of mechanical tactile stimuli delivered at the fingertips on SEEG contacts that were spatially clustered. The findings in this study suggest that evoking fingertip percepts through intracortical stimulation would require accessing the sulcal regions of S1. Additionally, it shows the potential of SEEG electrodes in providing the kind of somatosensory feedback most required for dexterous hand movement in sensorimotor BCIs.

We used neuroimaging tools designed by the Human Connectome Project to aid electrode implantation. Though the fMRI activation maps highlight the somatotopic subregion corresponding to the hand area in S1, the cortical areas of S1 are not clearly delineated. In this study, the intrinsic blood-oxygen-level-dependent (BOLD) point spread function at 3T, and the less well-defined finger task do not allow us to detect fMRI activation patterns of the distal (i.e., fingertip) vs proximal phalanges. However, previous studies with 7T fMRI have been able to localize representation of the fingertips in S1 (18). The group average cortical parcellations based on T1w/T2w-based myelin content and cortical thickness maps could effectively guide implantation of SEEG electrodes to target the finger representations in S1.

The well-known somatotopy of the human S1 represents the digits D1 (thumb) to D5 (little finger) in lateral-to-medial succession from the lateral border of the upper extremity subregion. Such somatotopic mapping was characteristically depicted in the task fMRI activations in area 3b in both participants (20, 21), notwithstanding the possibility of visually-driven contribution (52), while thenar (muscles under the base of the thumb) and palm sensation during finger tapping likely account for the activations observed in areas medial to the D5 representation. We interpret the overlapping activations from all fingers within the representation of D1 in area 1 as an inadvertent result of increased counter-balance pressure of the thumb pressing against the surface underneath, while the other fingers were lifted during finger tapping. Among the fingers, the thumb shows the most consistent and distinct representation in the expected somatotopic subregion. This is consistent with the relatively isolated anatomical structure of D1 from other fingers. The less consistent and focal representation of the other fingers are potentially due to a couple of reasons. First, unlike other elegant sensory task designs with dedicated tactile or electrical stimulation devices for each finger, sensory activations from common motor tasks are inherently imprecise despite best-efforts in task instruction and subject compliance. Second, the isolation of single digit motion is much more difficult for the rest of the digits than the thumb.

Sulcal stimulation in human S1 evoking fingertip percepts more often than gyral stimulation is in agreement with the imaging studies that have predominantly localized fingertip representation to the posterior wall of the central sulcus (area 3b) and occasionally at the crown of the postcentral gyrus (18–21, 28). The representation of fingertips might still extend into the posterior regions of the postcentral gyrus as shown by some imaging studies (25, 26, 30) and recent stimulation studies (13, 27). However, in our study, we had only two SEEG electrodes located deep in the postcentral sulcus in one participant that evoked percepts in the hand. None of the HD-ECoG electrodes located in the posterior regions of the postcentral gyrus evoked fingertip percepts. It might be the case that the fingertip representations on the postcentral gyrus are small and require extremely precise targeting while they are more extensive within the central sulcus and hence, easily accessible with SEEG electrodes.

In case of BCI research, both microelectrode arrays and ECoG grid electrodes have been used to provide artificial sensory feedback through intracortical or cortical surface stimulation, respectively. But these electrodes are restricted to stimulating the gyral areas of the cortex. Moreover, microelectrode array contacts tend to activate very closely spaced cortical locations resulting in the evoked percepts being restricted to relatively small anatomical regions, often spanning only a few phalanges over two or three fingers (11–13). Such high degree of anatomical overlap among the evoked percepts restricts the amount of sensory information that can be conveyed. Meanwhile, with their the larger size and inter-contact spacing, HD-ECoG electrodes elicit more diffuse sensory percepts (14, 16, 53). Additionally, with the capability to record or modulate the neural activity of neurons that lie within 1-2 mm below the cortical surface (54), these electrodes provide access to only the gyral surfaces of the cortex.

With the potential to reach sulcal areas of the cortex, SEEG electrodes provide a unique advantage of being able to evoke tactile sensations that are perceived to emanate from the fingers, particularly fingertips. Such percepts would be highly advantageous for providing a highly intuitive and useful sensory feedback while users of BCI handle objects. We demonstrate that SEEG electrodes can provide access to the sulcal areas and evoke highly focal sensory percepts at the fingertips. A recent study has reported being able to target fingertip representations on the postcentral gyrus using microelectrode arrays in a patient with SCI (13). However, such precise targeting was possible only after performing extensive intraoperative mapping of neural responses in S1 to peripheral stimulation of the fingertips relying on the relatively intact residual sensory pathways. Such an approach based on evoked responses in S1 might not be feasible in case of other potential users of BCI with more severe loss of function.

SEEG electrodes have recently been gaining favor for seizure onset localization as well as for mapping eloquent areas of the cortex in case of medically refractory epilepsy (32). In contrast to other intracranial electrodes that require burr holes or large craniotomies for implantation, SEEG electrodes can be implanted using a minimally invasive approach via a 1–2 mm craniostomy (31). The minimally invasive approach for their implantation reduces the risk of hemorrhage and infection to 1% and 0.8% respectively from that of 4% and 2.3% for subdural electrodes such as ECoG grids (38, 39).

In this study, all stimulation was done using a bipolar configuration involving adjacent electrodes. In case of HD-ECoG grids, with the electrodes arranged on the cortical surface, bipolar stimulation can potentially activate a wide area of the cortex evoking diffuse sensory percepts (55). Since SEEG electrodes can be inserted parallel to the orientation of the cortical columns and a contact diameter of only 0.8 mm, a bipolar stimulation configuration can potentially restrict stimulation to a couple of columns. This translates to the activation of neural circuits located in the receptive field of a highly specific area of the body, which in case of S1 stimulation, could result in highly focal sensory percepts. However, stimulating deeper structures could result in activating white matter tracts which could be counterproductive in terms of achieving focal sensory percepts. It has been shown that stimulating beyond 7 mm from the cortical surface does not trigger functional effects (36). Currently, the large inter-contact distances in SEEG leads used in this study (4.4 mm center to center) is potentially a limiting factor in achieving more focal percepts. Future studies should explore denser and smaller SEEG electrodes that could restrict the effective volume of cortical tissue that is activated and thus, evoke more focal percepts. However, higher current density and the potential tissue damage are factors that will have to be considered as well.

The MCC (repeatability metric) of recorded neural activity during mechanical tactile stimulation of the fingertips highlighted SEEG contacts that were either identical or adjacent to those that evoked percepts in the hand. This data further supports that the sulcal locations identified during electrical stimulation are indeed related to and important in tactile sensory restoration. This could also potentially provide a safer way to map eloquent cortex avoiding direct electrical stimulation which could trigger after-discharges or seizure activity (56) as well as for recording task-related, highly relevant neural activity for BCI applications.

Thus, we have shown that the representation of fingertips is readily accessible on the posterior wall of the central sulcus using SEEG electrodes. This suggests that SEEG electrodes offer a highly viable alternative to ECoG and microelectrode arrays for restoring somatosensation using intracortical stimulation. Future technical developments that will allow tightly spaced electrodes are important for greater success and efficacy. A recent review explored the potential of SEEG electrodes in BCI applications for decoding intended movement (57). Combined with our findings on SEEG electrodes being able to provide highly focal and relevant somatosensory feedback, we venture that sulcal stimulation via SEEG electrodes can potentially become an established approach for sensory restoration in closed-loop BCI applications.

## Data Availability

Data from the publication is available from the corresponding author upon request.

## Acknowledgements

We would like to thank our participants for their extraordinary commitment to this study, their patience with the experiments, and the deep insights provided by them, the clinicians and researchers at Feinstein Institutes for Medical Research. We would also like to thank Kevin Tracy, MD for providing critical inputs on the manuscript.

## Author Contributions

SB, JLH, CEB and ADM designed the study. JWK and JX designed and performed the fMRI procedures at Icahn School of Medicine at Mount Sinai and analyzed the data. ADM performed the SEEG leads and HD-ECoG grid implantations. NM and EE digitized and co-registered the electrode locations. SC, SB, NAB, RR and CEB performed all the experiments. SC and CEB analyzed data from these experiments. MFG and JX provided key insights into cortical anatomy, help using the workbench software and generating relevant figures for the manuscript. All authors contributed towards interpreting the results of the experiments. SC and CEB finished the initial draft of the paper and all authors provided critical review, edits and approval of the final manuscript.

## Supplementary Figures

**Fig. S1.**
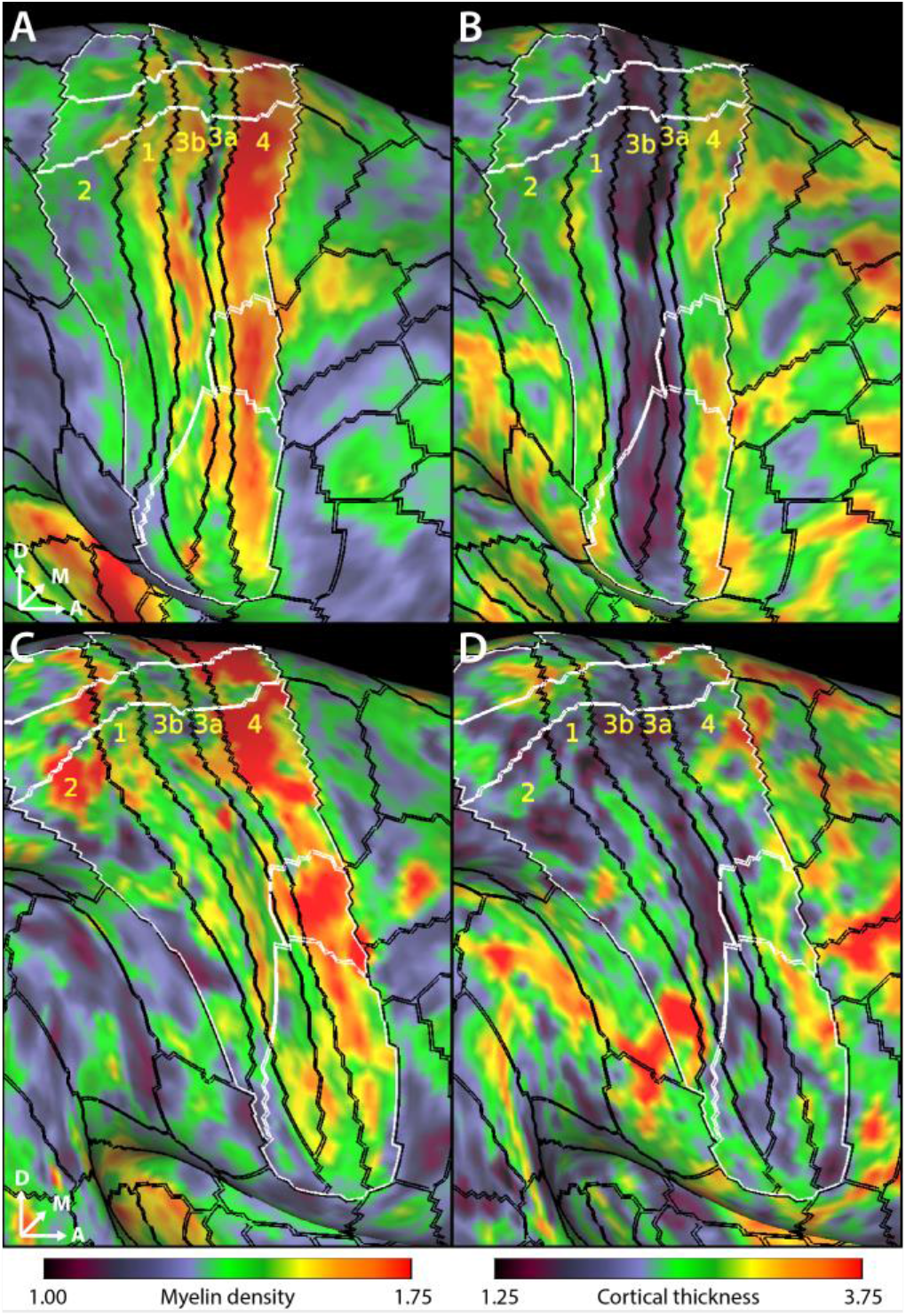
Group cortical areal boundaries line up with individual myelin and cortical thickness maps. **A**. Color map shows the T1w/T2w-based myelin map for participant 1 as per color bar at bottom. **B**. Color map shows the cortical thickness for participant 1 as per color bar at bottom. **C**. Color map shows the T1w/T2w-based myelin map for participant 2. As per color bar below. **D**. Color map shows the cortical thickness for participant 2 as per color bar below. The black lines overlaid on the cortical surface delineate the cortical areal boundaries outside of sensorimotor cortex and sensorimotor subregional boundaries inside sensorimotor cortex as derived from the HCP S1200 group average parcellation (40) based on myelin and cortical thickness maps and are labeled in yellow letters. The white lines demarcate the sensorimotor subregions based on resting state and task-based functional MRI. The sensorimotor subareas are denoted by the intersection of the black and white boundaries.

